# *SCENTinel 1.0*: development of a rapid test to screen for smell loss

**DOI:** 10.1101/2020.12.10.20244301

**Authors:** Valentina Parma, Mackenzie E. Hannum, Maureen O’Leary, Robert Pellegrino, Nancy E. Rawson, Danielle R. Reed, Pamela H. Dalton

**Affiliations:** Department of Psychology, Temple University, Philadelphia (PA); Monell Chemical Senses Center, Philadelphia (PA)

**Author notes:** **Corresponding authors:** Valentina Parma, PhD, Department of Psychology, Temple University, Weiss Hall, Room 874, 1701 North 13th Street, Philadelphia, PA 19122, Pamela H. Dalton, PhD, MPH, Monell Chemical Senses Center, 3500 Market Street, Philadelphia, PA 19104.

## Abstract

**Background:** Commercially available smell tests are primarily used in research or in-depth clinical evaluations, but are too costly and lengthy for population surveillance in health emergencies like COVID-19. We developed the *SCENTinel 1*.*0* test which rapidly evaluates three olfactory functions (detection, intensity, and identification). We tested whether self-administering the *SCENTinel 1*.*0* test discriminates between individuals with smell loss or average smell ability (normosmics), and provides comparable performance as the validated and standardized NIH Toolbox^®^ Odor Identification Test in normosmics.

**Methods:** Using Bayesian linear models and prognostic classification algorithms, we compared the *SCENTinel 1*.*0* performance of a group of self-reported anosmics (N=111, 47±13yo, F=71%,) and normosmics (N=154, 47±14yo, F=74%), as well as individuals reporting other smell disorders (e.g., hyposmia, parosmia; N=42, 55±10yo, F=67%).

**Results:** Ninety-four percent of normosmics met our *SCENTinel 1*.*0* accuracy criteria, while only 10% of anosmics and 64% of individuals with other smell disorders did. Overall performance on *SCENTinel 1*.*0* predicted belonging to the normosmic group better than identification or detection alone (vs. anosmic: AUC=0.95, Sensitivity=0.72, Specificity=0.94). Odor intensity provided the best single-feature predictor to classify normosmics. Among normosmics, 92% met the accuracy criteria at both *SCENTinel 1*.*0* and the NIH Toolbox^®^ Odor Identification Test.

**Conclusions:** *SCENTinel 1*.*0* is a practical test able to discriminate individuals with smell loss and is likely to be useful in many clinical situations, including COVID-19 symptom screening.

## Introduction

The COVID-19 pandemic has shown us how vulnerable we are to diseases that find an entry point in the olfactory system (Brann et al., 2020; Cooper et al., 2020; Pellegrino et al., 2020; Rodriguez et al., 2020). Despite how common the sudden onset of smell loss is in people with COVID-19 (Menni et al., 2020; Yan et al., 2020; Roland et al., 2020), the sense of smell is rarely evaluated in routine medical care, an omission which can have significant negative clinical implications (i.e., missed early identification of neurodegenerative disorders, lack of development of treatment options (Neuland et al., 2011; Croy et al., 2014; Boesveldt et al., 2017; Erskine and Philpott, 2020). The failure to see the mainstream clinical potential of evaluating the sense of smell is due to both theoretical and practical factors. Smell may be viewed as unimportant or as a vestigial sense despite a wealth of evidence to the contrary (McGann, 2017). As a result, olfactory function is rarely assessed until an individual experiences a significant - often complete - smell loss, and the lack of both primary and specialty physicians able to evaluate ‘normal’ olfaction apart from questionnaires is widespread, leaving the diagnosis of olfactory loss to a few specific specialties. Such shortage of widespread olfactory assessments spanning across the lifespan likely results in the underestimation of the true prevalence of smell loss in the general population. However, as COVID-19 revealed, such routine and rapid smell tests for population surveillance are needed. A recent meta-analysis highlights the sensitivity of direct measures of smell compared with self-reports (Hannum et al. 2020). Unless it is measured directly, many people do not realize their sense of smell is partially reduced, which might at least contribute to explaining why three-quarters or more of people with COVID-19 self-report no symptoms at all (Letizia et al., 2020; Petersen and Phillips, 2020). However, infection with COVID-19 is not the only cause of olfactory disorders. Indeed, anosmia (total loss of smell) and hyposmia (decreased ability to smell) can be caused by many respiratory viruses, including the common cold (Temmel et al., 2002; Pellegrino et al., 2017; Cavazzana et al., 2018), as well as sinonasal disease, neurodegenerative disorders and head trauma among others (Nordin and Brämerson, 2008; Doty, 2017; Hummel et al., 2017; Dalton, 2004; Damm et al., 2002).

### Current smell tests do not meet requirements for population surveillance

There are a number of commercially available and validated smell tests (Doty et al., 1984, 1996; Hummel et al., 1997; Choudhury et al., 2003; Jackman and Doty, 2005; Dalton et al., 2013; Rawal et al., 2015; Liu et al., 2020; Duff et al., 2002; Croy et al., 2015; Kondo et al., 1998). These tests are suitable for research and in-depth clinical testing yet they do not meet the scientific and practical needs for population surveillance, e.g., speed and low cost. There are several ways to measure olfaction, to see if a person can detect/discriminate the presence of an odorant or can correctly identify the odorant. Rating the intensity of an odorant offers an additional option, which while used in research, is not a component that is assessed in commercial smell tests. Most existing smell tests only include a single olfactory task, odor identification (Duff et al., 2002; Jackman and Doty, 2005; Dalton et al., 2013; Rawal et al., 2015; Doty et al., 1984). Although the most popular, odor identification is also the most sensitive among olfactory skills to cognitive deficits (e.g., verbal memory impairment, Wilson et al., 2006; Hedner et al., 2010), which can result in impaired performance for non-sensory reasons. Odor identification alone may fail to detect the reduction in intensity (especially among young people, who contrary to a more elderly population, may have lost much ability to smell, but nevertheless retain enough ability to guess the odorant’s quality). Additionally, odor intensity, even when self-reported, has proven to be the most predictive symptom indicator of a COVID-19 diagnosis (Gerkin et al., 2020). Indeed, either an odor detection, discrimination, or identification test can reveal whether an individual suffers from functional anosmia. Yet, if their sense of smell is only partially diminished (hyposmia) or distorted (parosmia), testing different smell functions will reveal divergent results. For example, a person with hyposmia may detect and discriminate a target odor depending on concentration and if so may identify an odor’s quality. However, a person with parosmia may detect and discriminate an odor but fail to identify it. Indeed, measuring different olfactory functions reveal response patterns commonly associated with different etiologies (Whitcroft et al., 2017). Therefore, there is a need to develop a smell test that rapidly assesses multiple olfactory functions in order to provide an assessment of smell loss which can be optimized for routine use and population surveillance.

### Large scale deployment of smell test for population surveillance

At least six considerations are important for large scale deployment of a smell test: (a) fast execution and administration without trained personnel, (b) use of easily identifiable odorants, (c) several test versions to allow for people to take the test frequently, (d) uniform delivery of odorants across sessions, (e) protection from physical contamination while taking the test, and (f) the correct answers must not be not easy to guess. **Speed** is important because smell testing especially for population surveillance must be fast, e.g., for building admittance. **Odorant choice** is important because the odorants must be familiar within the cultural or geographic context where the test is used, to minimize misattributions that do not depend on the ability to smell (Rabin and Cain, 1984; Ayabe-Kanamura et al., 1998). Odorants should not have a pungent component due to trigeminal activation (e.g., mint and cinnamon) because they can be detected by anosmic individuals (Laska et al., 1997). The **number of odorants** is important because the test could be repeatedly taken (e.g., each day for several weeks), and it should include enough odorants so that people do not give rote answers. **Uniformity** in how the odorant is delivered is important (e.g., odorant pens) and they should be easily accessed without tools (e.g., coins which are often used for scratch-and-sniff) and without introducing new sources of variation (e.g., unequal scratching when releasing the odorant). Avoiding **physical contamination** is important and participants cannot share the same olfactory stimulus (e.g., single-use, disposable tests to reduce the transfer of potential pathogens from nose to hand). Finally, the test must be **robust against guessing**.

### The *SCENTinel 1*.*0 Smell Test*

To meet the six above-mentioned criteria, we designed *SCENTinel 1*.*0* (a portmanteau of “scent” and “sentinel”). The self-administered test rapidly assesses three components of olfactory function: odor detection, intensity, and identification. To assess the performance of *SCENTinel 1*.*0*, we have conducted a quantitative cross-sectional study. The objective of the present research was to i) evaluate the ability of *SCENTinel 1*.*0* to discriminate between individuals suffering from anosmia and normosmics, as well as ii) determine the performance of *SCENTinel 1*.*0* compared to a validated and standardized gold-standard smell test (i.e., NIH Toolbox^®^ Odor Identification Test) in a normosmic group.

We hypothesize that

i. Normosmics will meet the *SCENTinel 1*.*0* accuracy criteria at a higher rate than the anosmic group and individuals with other olfactory disorders;
ii. In the normosmic group, overall *SCENTinel 1*.*0* performance is comparable to the performance on the NIH Toolbox^®^ Odor Identification Test.

## Methods

The materials, procedures, hypotheses and pre-analysis plan were all pre-registered and are available in the Open Science Framework Repository (OSF, Parma et al., 2020). Additional analyses (i.e., prognostic classification analyses) are marked as exploratory.

### Components of *SCENTinel 1*.*0*

*SCENTinel 1*.*0* is rapid and less expensive than the current commercially available validated smell tests. It measures odor detection, intensity and identification based on evaluation of a single odorant. Here, we assessed *SCENTinel 1*.*0*, which version used a flower odor (Givaudan, perfume compound with 2-phenylethanol CAS-No. 60-12-8 as the main component). *SCENTinel 1*.*0* is comprised of three patches, created with the Lift’nSmell^®^ technology (Scentisphere, Carmel, NY), glued to a card via an adhesive, only one of which contains an odorant. This technology prevents cross-contamination of odor to the ‘blank’ patches on the same card (imperative for an accurate odor detection test), promotes the standardization of odor delivery across cards and odors (imperative for an accurate odor intensity test), and limits residual odor in the air after the test (imperative for accurate odor identification).

To complete the fulfillment of the scientific and practical criteria above, *SCENTinel 1*.*0* includes olfactory functions that can be objectively assessed to yield a falsification metric and enable the ability to calculate the probability of meeting the test’s accuracy criteria in the absence of smell ability. The **odor detection subtest** has a guessing probability of 33%. The **odor intensity subtest** relies on the subjective experience of the participant and cannot be directly falsified. Intensity was included because a cutoff rating (i.e., <20 on a 1-100 scale; Gerkin et al., 2020) can be predetermined to signal smell loss for an odorant generally perceived as moderate to strong, and useful for tracking an individual’s smell function over time (i.e., identifying changes with repeated testing). The **odor identification subtest** comprises two possibilities: the first attempt, which is a 4-alternative forced choice task with guessing probability of 25% and a second attempt for those who failed the first attempt, which is a 3-alternative forced choice task with guessing probability of 33%. To allow for comparability, we used the NIH Toolbox® Odor Identification Test flower distractors (Dalton et al., 2013). For a full report on the instructions see the Procedures section and for the possible response patterns and the *SCENTinel 1*.*0* accuracy matrix, please refer to **Table 1**.

**Table 1.** *SCENTinel 1*.*0* accuracy matrix: potential response patterns and guessing probabilities.

| Response Pattern # | Detection | Intensity (range 1-100) | Identification |  | P(ch) |
| --- | --- | --- | --- | --- | --- |
|  |  |  | First attempt | Second attempt |  |
| 1 | Correct | > 21 | Correct | NA | 0.07 |
| 2 | Correct | ≤ 20 | Correct | NA | 0.02 |
| 3 | Correct | > 21 | Incorrect | Correct | 0.07 |
| 4 | Correct | ≤ 20 | Incorrect | Correct | 0.02 |
| 5 | Correct | > 21 | Incorrect | Incorrect | 0.13 |
| 6 | Correct | ≤ 20 | Incorrect | Incorrect | 0.03 |
| 7 | Incorrect | > 21 | Correct | NA | 0.13 |
| 8 | Incorrect | ≤ 20 | Correct | NA | 0.03 |
| 9 | Incorrect | > 21 | Incorrect | Correct | 0.13 |
| 10 | Incorrect | ≤ 20 | Incorrect | Correct | 0.03 |
| 11 | Incorrect | > 21 | Incorrect | Incorrect | 0.26 |
| 12 | Incorrect | ≤ 20 | Incorrect | Incorrect | 0.07 |

Grey shaded row: accurate response patterns; # =response pattern number. Detection is by a triangle test. "First attempt" is a four-alternative forced-choice. "Second attempt" is a three-alternative forced choice. P(ch)=probability of an outcome by chance.

### Participants

Eligible participants were recruited via an electronic flyer distributed through the Monell Newsletter, allowing the enrollment of normosmic subscribers and subscribers with different forms of smell loss (**Figure 1**).

**Figure 1.**
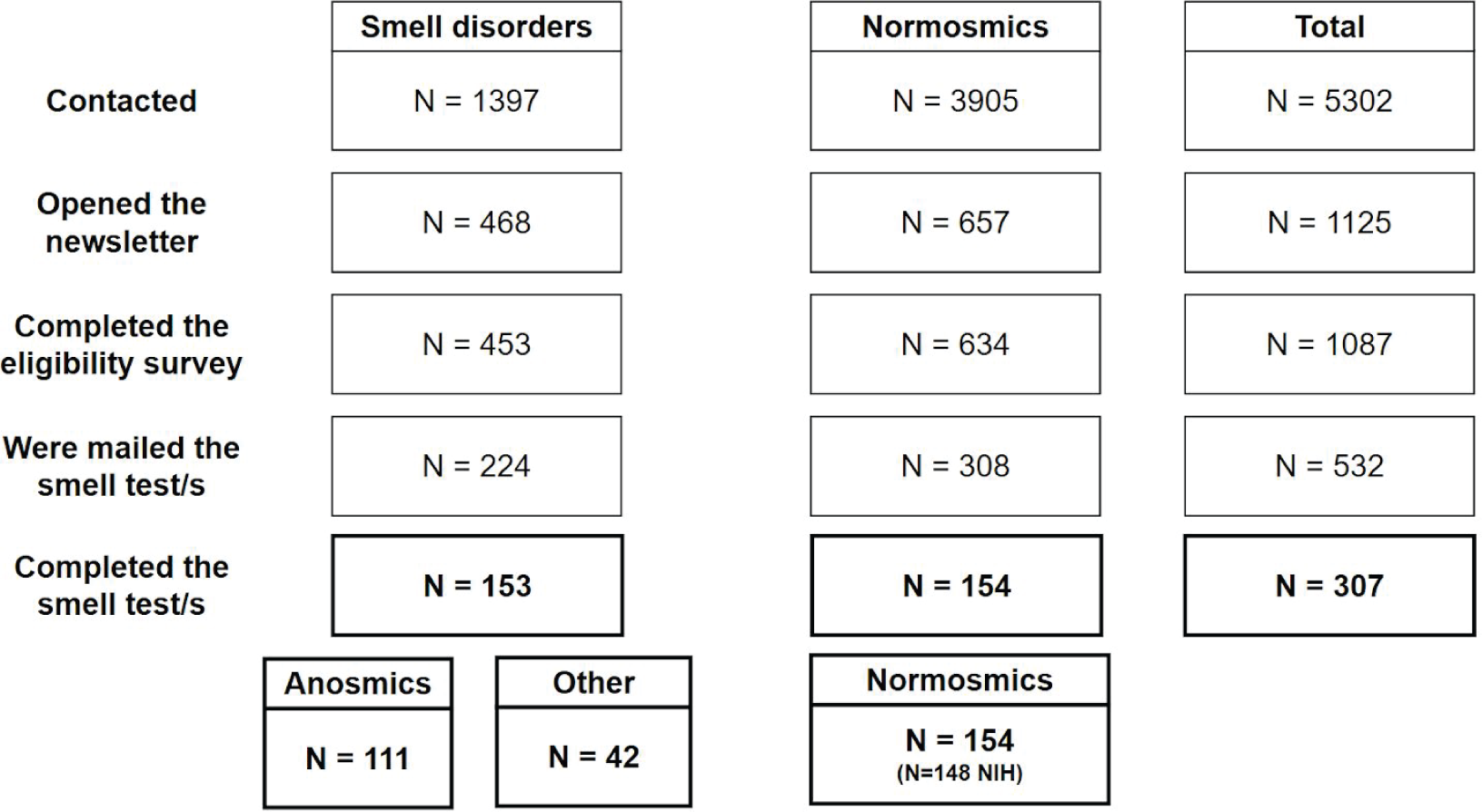
Diagram showing the sample description by group. Other: Individuals who self-reported other smell disorders; NIH = Normosmics who completed the NIH Toolbox® Odor Identification Test.

Volunteers completed an eligibility survey (**Appendix I**) in which they reported their age (inclusion criteria: 18-75 years old, 257 excluded), whether they had access to a smart device (phone or tablet) or a computer (6 excluded), and whether they were currently residing in the United States (121 excluded).

A total of 532 *SCENTinel 1*.*0* tests were distributed by mail on a first-come, first-served basis; 308 participants reporting no history of smell problems received one *SCENTinel 1*.*0* test, and one NIH Toolbox® Odor Identification Test (Dalton et al., 2013). Participants with pre-existing forms of smell loss (N = 224) received one *SCENTinel 1*.*0* test, and were not asked to complete the NIH Toolbox^®^ Odor Identification Test to limit the emotional burden generated by participating in smell tasks. Participants were also invited to take *SCENTinel 1*.*0* (and the NIH Toolbox^®^ Odor Identification Test, if provided) on the same day they were scheduled to have a COVID-19 PCR test. We then asked them to report the results of the COVID-19 PCR test via survey when the outcome was known. Only 3 participants took the smell test/s and the COVID-19 PCR test on the same day, thus given the low numerosity we excluded this variable from the analyses. The completion rate of those who were sent a smell test was 58%, with a final sample size of participants who consented and participated in the study including 154 normosmic adults, 111 anosmics and 42 participants with other smell disorders [fluctuations (N=5); hyposmia (N =23), parosmia (N=5), other (N=4), COVID-related smell loss (N=3)]. Given the low numerosity in each subgroup of the ‘other smell disorders’ variable, no separate statistical analyses were performed on this factor. Please refer to **Table 2** for a description of the demographics of the sample. One-hundred and forty-eight normosmic participants also completed the NIH Toolbox^®^ Odor Identification Test.

**Table 2.** Description of the final sample who completed *SCENTinel 1*.*0*.

|  |  | **Anosmic** | **Other smell disorders** | **Normosmic** |
| --- | --- | --- | --- | --- |
| **Age (yo)** | **mean±sd** | 47±13 | 55±10 | 47±14 |
|  | **range** | 19-72 | 32-69 | 20-74 |
| **Sex** | **F (%)** | 79 (71%) | 28 (67%) | 114 (74%) |
|  | **M (%)** | 32 (29%) | 14 (33%) | 40 (26%) |
|  | **Prefer not to say (%)** | 0 | 0 | 0 |
| **Race/**<br>**Ethnicity** | **Asian (%)** | 3 (3%) | 0 | 12 (8%) |
|  | **Black (%)** | 3 (3%) | 1 (2%) | 5 (3%) |
|  | **Hispanic (%)** | 2 (2%) | 0 | 6 (4%) |
|  | **Native Hawaiian (%)** | 0 | 0 | 1 (1%) |
|  | **Other (%)** | 2 (2%) | 2 (5%) | 1 (1%) |
|  | **Prefer not to say (%)** | 1 (1%) | 1 (2%) | 1 (1%) |
|  | **White (%)** | 100 (90%) | 38 (90%) | 128 (83%) |
|  | **N total** | **111** | **42** | **154** |

yo = years old; sd = standard deviation.

### Procedures

The study started on September 4th, 2020 and was completed by September 15th, 2020. During this time, participants were contacted via the Monell newsletter mail list, and completed a 10-question online eligibility survey (**Appendix I**). Subscribers to the Monell newsletter mail list include volunteer leadership, academic, industry and organizational partners, donors, individuals with health-related interest in the research conducted at Monell, and individuals who have attended Monell events. In that process, participants provided their electronic consent via REDCap. The study was approved using a waiver of documentation of consent by the Institutional Review Board of the University of Pennsylvania (#821887). If they were not eligible or if they responded after the target number of participants had been enrolled, they were thanked and informed that they would not be enrolled in the study (N = 555, **Figure 1**). If, on the contrary, they were deemed to be eligible and tests were still available, they received one or two smell tests via mail depending on their anosmic/normosmic self-report status. Once participants received the test, they were instructed to complete them within the next 14 days. Participants used a QR-code or a web address to access the REDCap survey (Harris et al., 2019) used to record self-reports on demographic data (age, gender, ethnicity), pre-existing smell and taste loss, as well as instructions to complete *SCENTinel 1*.*0* and the NIH Toolbox^®^ Odor Identification Test, if provided. To complete *SCENTinel 1*.*0*, the instructions were to consecutively open one odor patch at a time, smell each patch and reseal; (a) choose the patch with the strongest odor-; (b) rate the intensity of the odor on a visual analog scale from 0 (no smell) to 100 (very strong smell); and (c) select the best verbal and visual label for the odor among four options provided. If the participant gave an incorrect response to (c), they were instructed to try again to identify the odor, this time among the three remaining options. No additional feedback was provided on the accuracy of the odor identification after the second attempt. The group of participants who also completed the NIH Toolbox^®^ Odor Identification Test (normosmics) was instructed to scratch and sniff each of the 9 odors included in the NIH Toolbox^®^ Odor Identification Test and identify among 4 visual and verbal options which one corresponded to the odor smelled.The test consisted of nine odorants. Subsequently, the participants completing the NIH Toolbox^®^ Odor Identification Test could opt in to answer questions regarding their health status, with particular reference to COVID-19 and other respiratory illnesses. The results to those questions are irrelevant to the main hypotheses of this study and will be reported in a separate, future manuscript. Although no formal data were collected on the completion time of *SCENTinel 1*.*0* in the present sample, pilot participants (N = 10, 9 F, 27-65 years old) reported that the test takes ∼2 minutes to complete when including the demographic questions and <1 minute to complete the *SCENTinel 1*.*0* subtests.

### Statistical analyses

This cross sectional design includes a between-subject factor “smell ability” (anosmic, other smell disorders, and normosmics) and within-subject factors, namely meeting the accuracy criteria within each subtest of *SCENTinel 1*.*0* (odor detection, intensity, identification) as well as the *SCENTinel 1*.*0* overall accuracy criteria (**Table 1**), and the scores at the single items and the total score at the NIH Toolbox^®^ Odor Identification Test.

Each *SCENTinel 1*.*0* subtest returns one of the following responses: odor detection accuracy (correct/incorrect); odor intensity, (above/below a cut off of 20) and odor identification among 4 given options (correct/incorrect), and if the first response is incorrect, among the three remaining options (correct/incorrect). The NIH Toolbox® Odor Identification Test returns two scores: the official scoring [anosmia = ≤3; hyposmia = 4-6; normosmia ≥ 7 (Dalton et al., 2013)] and a binarized version of the official score to enable direct comparison with the *SCENTinel 1*.*0* accuracy criteria (anosmia = ≤ 4; normosmia ≥ 5). This latter has been used in the present analyses.

We used a Sequential Bayes Factor design (SBFD) with maximal N, as suggested by Schönbrodt et al. (2017). This maximizes the probability of obtaining the desired level of evidence and a low probability of obtaining misleading evidence. Additionally, this SBFD design requires on average half the sample size compared to the optimal null hypothesis testing fixed-n design, with comparable error rates (Schönbrodt et al., 2017). The desired grade of relative evidence for the alternative vs. the null (BF_10_) hypothesis is set at BF_10_ > 6 (moderate evidence) for H_1_ and BF_01_ > 3 for H_0_ (anecdotal evidence). Based on a conservative Cohen’s D = 0.5, we have specified a minimum sample size per group of n_0_=43. Once n_0_ is reached, the BF will be computed on the existing data. BF computation will continue after every participant is added (in the smallest or slowest accumulating group at that time) until the thresholds of H_1_ or H_0_ are reached, at which point sampling will cease. The main driver of the stopping rule is, however, a time limit (September 15th). To test our hypotheses and explore covariate effects (age, sex, ethnicity) we employed Bayesian linear mixed models using the *BayesFactor* package (Morey et al., 2018) in the R Environment for Statistical Computing (R Core, 2020). For analyses, given the unequal distribution of the data across categories in the ethnicity variable, we have binarized the responses as White/Non-white. To assess the differences in accuracy among tests and subtests, we have employed Bayesian and parametric tests for equality of proportions with or without continuity correction.

In addition to the pre-registered analyses, we have applied machine learning prognostic classification algorithms to confirm the ability of *SCENTinel 1*.*0* to discriminate anosmics and normosmics, as well as individuals with other smell disorders. We removed the second trial of *SCENTinel 1*.*0’s* odor identification from the classification, given the high number of missing values and the challenges of imputation in those conditions. No imputation procedure was then required for the rest of the database. A one-hot encoding was applied to all categorical variables (sex and ethnicity) to produce binary indicators of category membership. Model quality was measured using receiver operating characteristic (ROC) area under the curve (AUC). We also report specificity, sensitivity, positive predictive value (PPV) and negative predictive value (NPV) based on the model that optimizes classification on unseen data among random forest, linear and radial small vector machine (SMV), regularized linear regression (Elastic net) and linear discriminant analysis (LDA). Cross-validation (number = 10, repeat = 5) was performed on the training set (80% of the sample) while validation was completed on the remaining, withheld data (20%). The model that provided the best classification AUC between anosmic and normosmic on the withheld data was LDA, which we report and discuss in the main text. The data and analysis script are available in the Supplementary material and will be publicly available on GitHub [link] upon publication.

## Results

### *SCENTinel 1*.*0* discriminates anosmics from normosmics

As expected, only a small group of anosmics (N = 11, 10%) met the accuracy criteria for *SCENTinel 1*.*0*. On the contrary, the majority of individuals with other smell disorders (N = 27, 64%), as well as the vast majority of normosmics met the accuracy criteria for *SCENTinel 1*.*0* (N = 145, 94%). As reported in **Figure 2** and **Table S1**, participants from the three groups primarily used different response patterns to complete *SCENTinel 1*.*0*. In the anosmic group, 23% of participants failed to meet the accuracy criteria for any of the subtests, 41% for two subtests, and only 11% failed to meet the accuracy criterion for odor intensity (i.e., reported intensity above 20/100). In the other smell disorders group, 17% of participants failed to meet the accuracy criterion for odor intensity, 17% for two out of the three subtests and only 2% for all three subtests.

**Figure 2.**
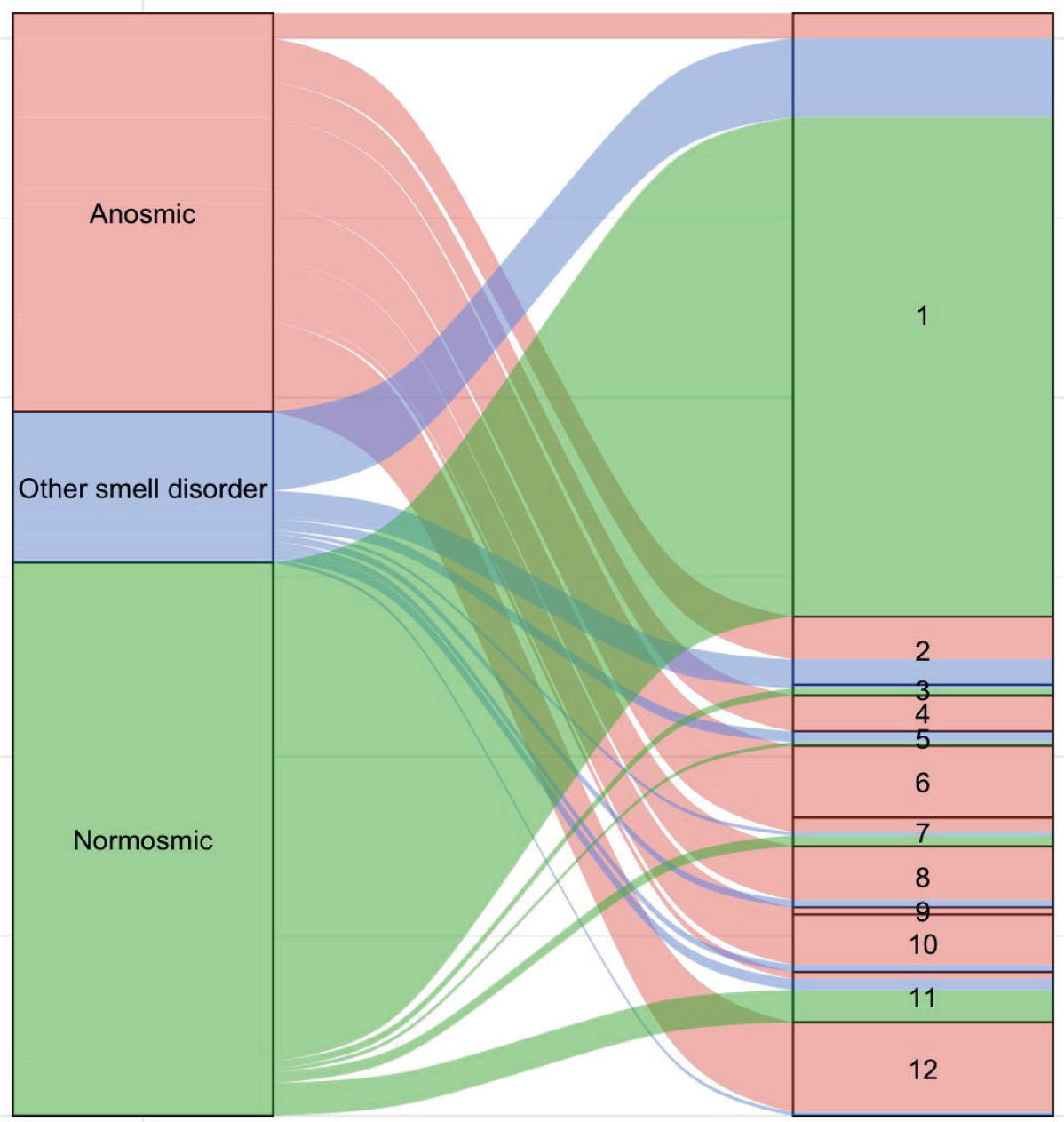
*SCENTinel 1*.*0* response patterns used by smell group (Anosmics = red; Other smell disorders = blue; Normomics = green). Response patterns 1, 3, 5, 7 indicate that participants met *SCENTinel 1*.*0*’s accuracy criteria.

The combined accuracy at all three subtests significantly discriminated the performance across the three groups. In particular, in this sample odor intensity is the subtest that demonstrates a perfect ability to identify normosmia (**Table 3**), as 100% of participants reported an intensity rating over the cut-off of 20. The only subtest that does not significantly discriminate between the performance of the three groups is the second attempt at odor identification, which was only used by 32 participants across the three groups (**Table 3**). No effects of age, sex or ethnicity across groups were revealed for any of the *SCENTinel 1*.*0* subtests (**Table S2**). A marginally moderate effect of age can be found in the performance of the first identification subtest (BF_10_ = 3.11, **Table S2**).

**Table 3.** Number and percentage of participants that in each smell group meet the accuracy criteria for *SCENTinel 1*.*0*, along with group comparisons.

| Subtests | Anosmics |  | Other smell disorders |  | Normosmics |  | Total |  | BF<sub>10</sub> | Proportions |  |  |
| --- | --- | --- | --- | --- | --- | --- | --- | --- | --- | --- | --- | --- |
|  | N | % | N | % | N | % | N | % |  | X<sup>2</sup>, df = 2 | p | significant comparisons |
| Odor detection | 49 | 44 | 33 | 79 | 142 | 92 | 224 | 73 | 5.33e+16 ± 0.01% | 76.32 | < 0.001 | a,b,c |
| Odor intensity | 15 | 14 | 30 | 71 | 154 | 100 | 199 | 65 | 8.85e+74 ± 0% | 212.52 | < 0.001 | a,b,c |
| Odor identification #1 | 38 | 34 | 32 | 76 | 142 | 92 | 212 | 69 | 3.55e+24 ± 0.01% | 102.61 | < 0.001 | a,b,c |
| Odor identification #2 | 26 | 23 | 3 | 7 | 3 | 2 | 32 | 10 | 0.20 ± 0.03% | 0.84 | 0.66 |  |

BF = Bayes Factor for the model $\text{lmBF}(\text{subtest score} \sim \text{Group}, \text{data}, \text{whichRandom} = \text{"ID"})$ ; ± X%: error of the estimate; df= degrees of freedom, p = p-value, a= comparison anosmics vs. normosmics; b = anosmics vs. other smell disorders; c = other smell disorder vs. normosmics.

We then examined which classification algorithm would best predict group-belonging to a particular smell group. Results from a recursive feature selection indicated that five features (odor detection, intensity, identification, age and female sex) recurred across samples. These results were confirmed by several other algorithms (**Figure S1**). To assess whether *SCENTinel 1*.*0* subtests would be sufficient to discriminate between different groups we investigated their ROCs with providing the greatest discrimination accuracy (LDA). As depicted in **Figure 3**, discrimination across the three smell groups is possible. The overall *SCENTinel 1*.*0* performance discriminated between anosmics and normosmics with greater accuracy (AUC = 0.95) than any of the subtests alone (**Figure 3A**). The intensity subtest appears to be the single best discriminator between anosmics and normosmics (AUC = 0.94), followed by odor identification #1 (AUC = 0.84) and odor detection (AUC = 0.80). Similarly, *SCENTinel 1*.*0* is also able to discriminate between individuals with other smell disorders and normosmics (**Figure 3B**), as well as anosmics vs. individuals with other smell disorders (**Figure 3C**). In this latter comparison, AUC is greatly reduced (AUC = 0.77). As hypothesized, each *SCENTinel 1*.*0* subtest differently contributes to the classification of individual performance and the contribution of each subtest to the classification is related to current smell ability. All *SCENTinel 1*.*0* subtests discriminate above chance anosmics from normosmics, yet the overall *SCENTinel 1*.*0* performance does so with greater confidence (**Figure 3A**). Odor detection and intensity discriminate above chance individuals with other smell disorders from normosmics, but odor identification does not (**Figure 3B**). Only the overall *SCENTinel 1*.*0* score discriminates above chance the performance of anosmics from individuals with other smell disorders, whereas no single subtest is able to do so in isolation (**Figure 3C**).

**Figure 3.**
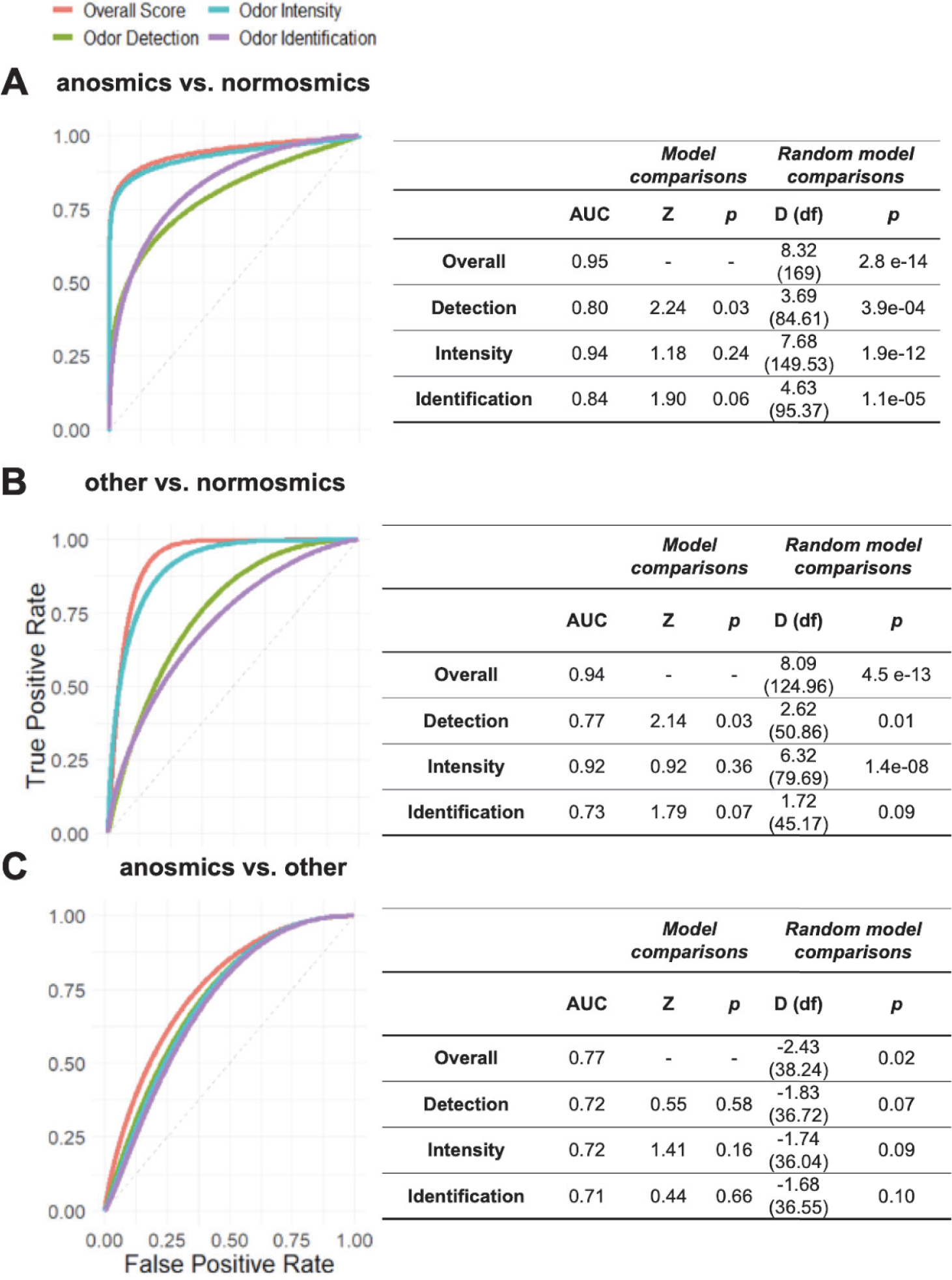
ROC curves and statistics on the overall *SCENTinel 1*.*0* score as well as single subtests across groups (anosmics vs. normosmics in panel **A**: other smell disorders vs. normosmics in panel **B** and anosmics vs. other smell disorders in panel **C**) based on the the Linear Discriminant Analysis algorithm. AUC = Area Under the Curve; p = p-value. D = DeLong’s test for two ROC curves; df = degrees of freedom.

### Performance on *SCENTinel 1*.*0* and on the NIH Toolbox^®^ Odor Identification Test is comparable in normosmics

Normosmics self-administered both *SCENTinel 1*.*0* and the NIH Toolbox^®^ Odor Identification Test in order to compare the performance of *SCENTinel 1*.*0* against a validated smell test. Results indicated that the performance on both tests is concordant when comparing performance on the flower odor identification, which was odor #9 in the NIH Toolbox^®^ Odor Identification Test (143/148, 97% participants correctly identified the flower odor) and the *SCENTinel 1*.*0* odor identification subtest (136/148, 92% accuracy in the first identification attempt). A 2-sample test for equality of proportions with continuity correction suggests the lack of statistical difference between the two test scores (X_2_ = 2.25, df = 1, p = 0.13). In 17/148 cases (12%) the NIH Toolbox^®^ Odor Identification Test and *SCENTinel 1*.*0* were discordant (**Figure 4A** and **4B**, red ribbons); specifically, in 12 cases the participant passed the NIH Toolbox^®^ Odor Identification Test but failed to meet the accuracy criteria for *SCENTinel 1*.*0* and in 5 cases the participant passed *SCENTinel 1*.*0* but failed the NIH Toolbox^®^ Odor Identification Test. When considering the full NIH Toolbox^®^ Odor Identification Test (9 items) and *SCENTinel 1*.*0* (detection, intensity and identification, both attempts) the accuracy converged: 92% of normosmics passed both tests. No effect of age (BF_10_ = 0.81 ±0.02%), sex (BF_10_ = 0.84 ±0.02%) or ethnicity (BF_10_ = 0.48 ±0.02%) was found for the perfomance on the NIH Toolbox^®^ Odor Identification Test.

**Figure 4.**
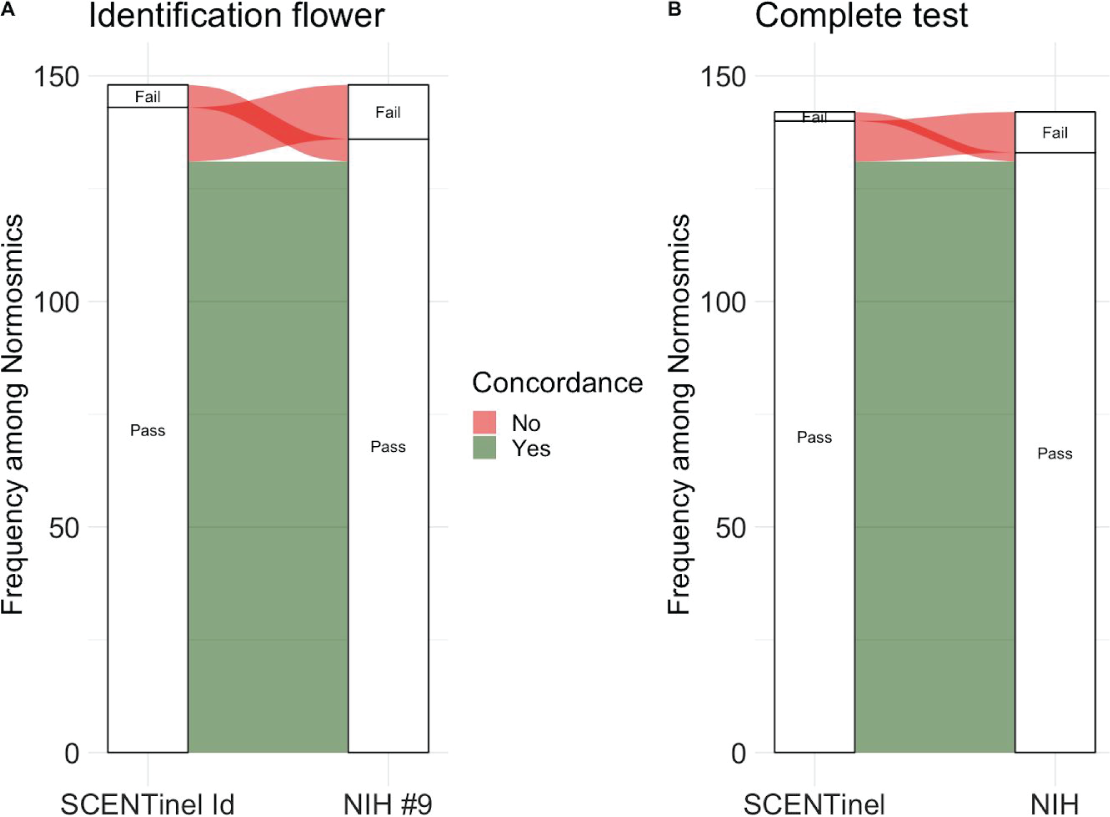
Concordance between *SCENTinel 1*.*0* and the NIH Toolbox^®^ Odor Identification Test in normomics. **A**. Concordance based on the odor identification performance at the flower odor. **B**. Concordance based on full completion of both smell tests.

## Discussion

The goal of the present study was twofold: to assess the *SCENTinel 1*.*0* performance to discriminate conditions of ongoing smell loss and normosmia and to compare the performance of *SCENTinel 1*.*0* to the NIH Toolbox^®^ Odor Identification Test, a validated and standardized gold-standard smell test. We hypothesized that normosmics will meet the *SCENTinel 1*.*0* accuracy criteria at a higher rate than both the anosmic group, as well as individuals with other olfactory disorders and that normosmics would similarly perform at *SCENTinel 1*.*0* and at the NIH Toolbox^®^ Odor Identification Test. Both of our main hypotheses were confirmed.

First, 94% of normosmics met the accuracy criteria, in contrast to the 64% of participants reporting other smell disorders and only 10% of participants reporting anosmia. The majority of participants with anosmia were not able to meet the accuracy criteria for two or three subtests, particularly the odor intensity subtest. In comparison, participants with other smell disorders failed to meet the accuracy criteria for two subtests (in particular, the odor intensity subtest) more often than the normosmic group. Normosmics met the accuracy criteria for all three subtests. The ability of the overall test to classify anosmics and normosmics based on performance is satisfactory (AUC = 0.95). The odor intensity subtest alone has also a similar classification ability (AUC = 0.94) but ratings of intensity can be intentionally misreported while the other subtests cannot. Odor identification represents the second best subtest in discriminating between normosmics and anosmics. Although this discrimination alone is less accurate compared to odor intensity, the odor identification subtest is an objective measure with a guessing probability of only 25% on the first attempt. Yet, the utility of the odor identification subtest is lost when discriminating normosics from those with other smell disorders. As anticipated, individuals suffering from hyposmia, which constitute the majority of the other smell disorders group, may be able to report on odor quality, but do not appropriately report its intensity. For individuals with parosmia, the performance could be different, yet these results cannot provide conclusive evidence given the low number of parosmic participants in this sample. Odor detection which offers a culturally-unbiased olfactory measure of olfactory performance (e.g., Doty et al., 2019), aids in concert with the other subtests, the discrimination of anosmia from other forms of smell loss.

Second, we established that the normosmics perform similarly well for both the *SCENTinel 1*.*0* with the NIH Toolbox^®^ Odor Identification Test. Ninety-two percent of participants were able to meet the accuracy criteria of both tests, and this figure increases to 97% when we consider the odor identification performance to a flower odor, which was the odorant tested here as well as the odor of item #9 in the NIH Toolbox^®^ Odor Identification Test.

We conclude that testing three olfactory functions with the goal of quickly detecting the presence of smell loss is possible and comparable to the performance on longer, validated and standardized tests, such as the NIH Toolbox^®^ Odor Identification Test. *SCENTinel 1*.*0* meets all the scientific and practical criteria outlined above for population surveillance based on smell testing. Specifically, it is structured to reduce the probability of passing the test by guessing alone and to be self-administered. Due to the Lift’nSmell^®^ technology, no tools are needed to complete the test (e.g., a coin) and the intensity of the odorant is not affected by participants’ behavior (e.g., scratching for scratch-and-sniff). Altogether, this test can be applied in a variety of contexts and for different purposes.

The findings presented here represent the first step of a broader research program that includes a full validation and normative study on the *SCENTinel 1*.*0* test. However, given the promising results and the urgent need to deploy all possible aids to control the spread of COVID-19, we report the data of this assessment. Presently, we have verified that *SCENTinel 1*.*0* is able to discriminate anosmics from normosmics, and among normosmics *SCENTinel 1*.*0* has been validated against the NIH Toolbox^®^ Odor Identification Test. Next, we are looking forward to extending the testing to the multiple *SCENTinel* versions that feature different odorants to assess whether performance is odor-invariant (Zernecke et al., 2010). We are currently developing eight versions of *SCENTinel 1*.*0* which use non-trigeminal odors, highly familiar to the US population, as indicated by published data from existing databases (Freiherr et al., 2012; Dalton et al., 2013).

Then, we will focus our efforts on clinically verifying the diagnosis of smell disorders in patients, since at present, participants self-reported normosmia and/or the ongoing presence of smell disorders, including anosmia. If verifying the clinical diagnosis is a necessary step from a research perspective, olfactory routine testing with large scale population deployment would likely lack this level of precision. It is therefore a very favorable result that *SCENTinel1*.*0* discriminates against different degrees of olfactory ability in individuals that lack an in depth research- or clinical-level investigation of their ability to smell. To this end, we intend to offer an analysis of the performance of *SCENTinel* across larger samples of individuals with hyposmia, parosmia, phantosmia, etc. to further the understanding of which olfactory functions have the most power in discriminating across smell disorders.

Other relevant individual variables that are known to affect olfactory performance include age, sex and ethnicity (Hedner et al., 2010; Menon et al., 2013; Sorokowski et al., 2019); though no differences were found in the present study. Although this initial study prominently featured women and White participants, unequally spread across different age groups, we aim at testing the performance of *SCENTinel 1*.*0* in more diverse groups to fully ascertain the effect of age, sex and ethnicity and to identify possible cross-cultural and genetic influences that may play a role in test perfomance. Additionally, the brevity of *SCENTinel 1*.*0* can facilitate its translation and widespread use across linguistic communities (e.g., native Spanish- and Chinese-speakers). Further monitoring intra-individual performance over time will not only provide a path to better understanding recovery from smell loss, but will also offer the opportunity to determine a lifespan surveillance approach to olfactory perception, following in the footsteps of the NIH Toolbox^®^ Odor Identification Test, which can be used from 3 years of age with minimal modifications and from age 10 in its full form (Dalton et al., 2013).

Altogether, our findings support the idea that *SCENTinel 1*.*0* represents a rapid, accurate, flexible, and cost-effective tool to deploy a smell test on large scale population surveillance efforts. The development of *SCENTinel 1*.*0* has been spurred by the new sudden loss of smell that characterizes COVID-19, including among nominally asymptomatic individuals, many of whom had smell loss but were not aware of before receiving an objective olfactory test (Gözen et al., 2020; Bhattacharjee et al., 2020). Yet, the large-scale availability of a validated rapid smell test can benefit not only health emergencies such as COVID-19, but also be used in early detection and monitoring of a variety of clinical conditions, including psychiatric, neurological and neurodegenerative disorders.

## Data Availability

The data and analysis script will be publicly available on GitHub upon publication.

## Acknowledgements

The authors wish to thank all study participants who enthusiastically and promptly completed the smell tests. We also would like to thank Jenifer Trachtman, Alyssa Wofford, Karen Kreeger, Steve Rowe, Dennis Coleman, Kathleen Bell, Matt Carr, Ronald Schwoyer, and Carol Christensen for their assistance with the development and distribution of the test to participants. We thank Mark Bernstein and Megan Hansen from Scentisphere Inc for the manufacture of the test. We thank Chris Maute for his contribution in science communication and we thank Riley Herriman and Heather Caviston for graphic design and artwork development.

## Funding

We acknowledge support from the National Institutes of Health as a part of the RADx-rad initiative (U01 DC019578 to PHD and VP), Monell Institutional Funds. Mackenzie Hannum and Robert Pellegrino are supported by NIH T32 funding (DC000014). This work was also supported by contributions for anosmia research from John Hickey, Philip Johnson, John Labows, Stephen Manheimer, Robert Margolskee, and hundreds of other individual donors. We thank Kerry and the Vernekoff Foundation for their support of this work.

## Conflict of interest

The authors declare no conflict of interest.

## Supplementary materials

**Table S1.**
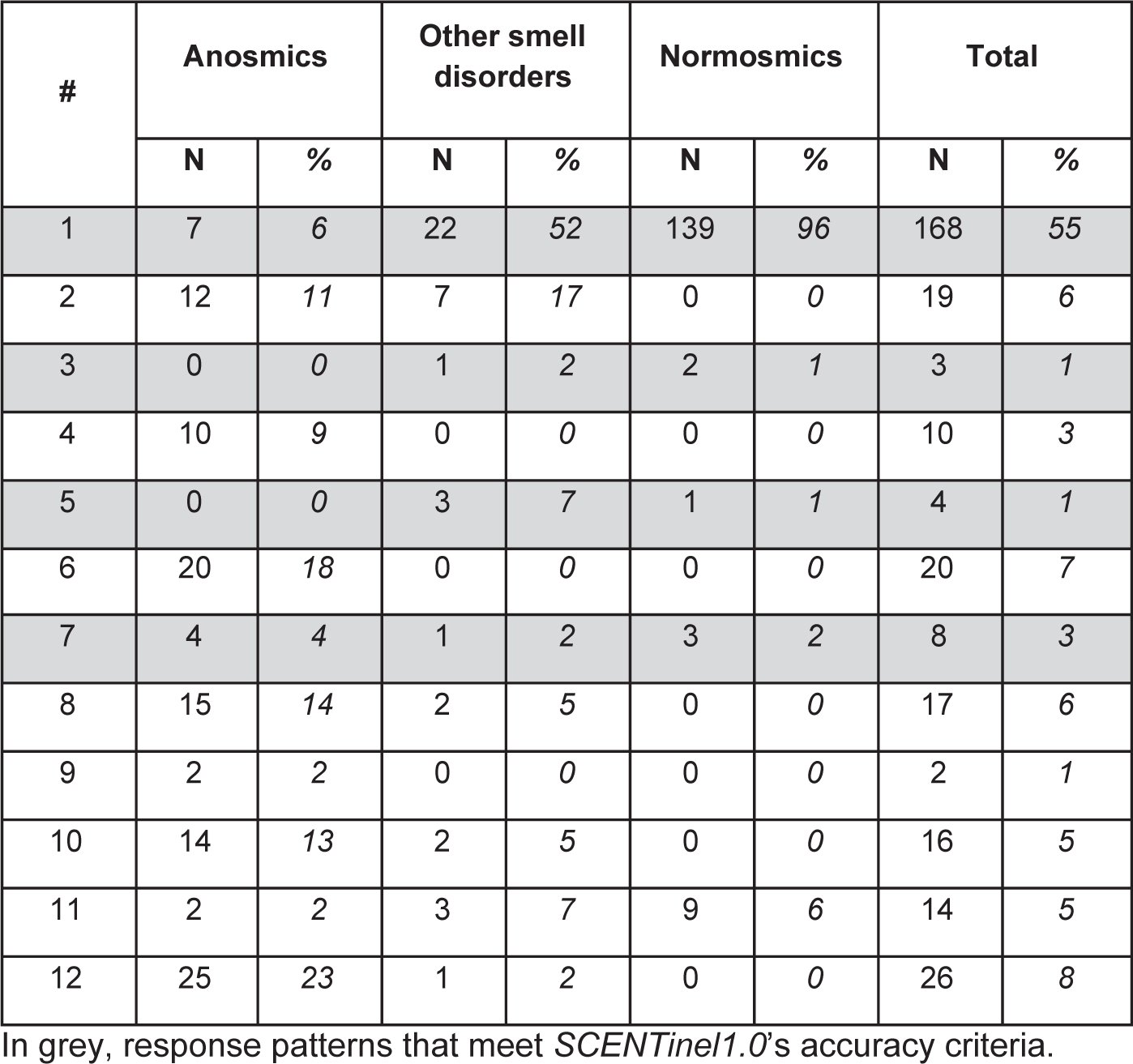
Number and percentage of participants who used a specific response pattern to complete *SCENTinel 1*.*0*

| # | Anosmics |  | Other smell disorders |  | Normosmics |  | Total |  |
| --- | --- | --- | --- | --- | --- | --- | --- | --- |
|  | N | % | N | % | N | % | N | % |
| 1 | 7 | 6 | 22 | 52 | 139 | 96 | 168 | 55 |
| 2 | 12 | 11 | 7 | 17 | 0 | 0 | 19 | 6 |
| 3 | 0 | 0 | 1 | 2 | 2 | 1 | 3 | 1 |
| 4 | 10 | 9 | 0 | 0 | 0 | 0 | 10 | 3 |
| 5 | 0 | 0 | 3 | 7 | 1 | 1 | 4 | 1 |
| 6 | 20 | 18 | 0 | 0 | 0 | 0 | 20 | 7 |
| 7 | 4 | 4 | 1 | 2 | 3 | 2 | 8 | 3 |
| 8 | 15 | 14 | 2 | 5 | 0 | 0 | 17 | 6 |
| 9 | 2 | 2 | 0 | 0 | 0 | 0 | 2 | 1 |
| 10 | 14 | 13 | 2 | 5 | 0 | 0 | 16 | 5 |
| 11 | 2 | 2 | 3 | 7 | 9 | 6 | 14 | 5 |
| 12 | 25 | 23 | 1 | 2 | 0 | 0 | 26 | 8 |

In grey, response patterns that meet *SCENTinel1.0*'s accuracy criteria.

**Table S2.** Assessment of the effect of demographic covariates on the *SCENTinel 1*.*0* subtests.

| Subtests | Demographic variable | BF10 |
| --- | --- | --- |
| Odor detection | Age | 0.18 ± 0.01% |
|  | Sex | 0.31 ± 1.37% |
|  | Ethnicity | 0.14 ± 0.97% |
| Odor intensity | Age | 0.33 ± 0.01% |
|  | Sex | 0.18 ± 1.22% |
|  | Ethnicity | 0.15 ± 1.05% |
| Odor identification #1 | Age | 3.11 ± 0.01% |
|  | Sex | 0.265 ± 2.32% |
|  | Ethnicity | 0.13 ± 2.22% |
| Odor identification #2 | Age | 0.11 ± 0.01% |
|  | Sex | 0.25 ± 1.19% |
|  | Ethnicity | 0.92 ± 7.37% |

BF<sub>Age</sub> = main effect model; BF<sub>Sex</sub> and BF<sub>Ethnicity</sub> = BF<sub>Sex(Ethnicity)+Group</sub>/ BF<sub>Group</sub>. The additive models represented the best models in all comparisons tested.

**Figure S1.**
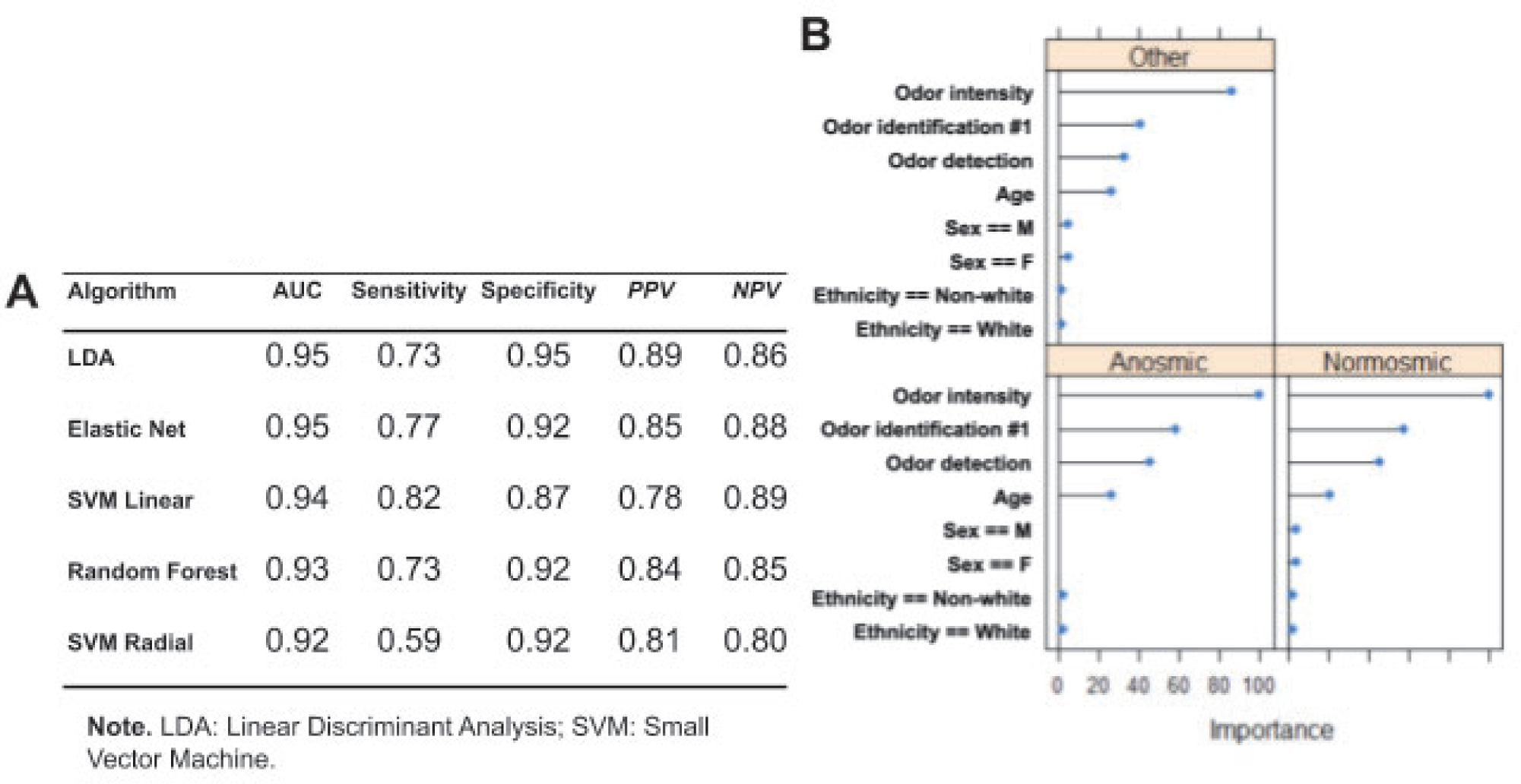
**A**. Prediction metrics for five algorithms used to predict multiclass group belonging (anosmics, other smell disorders, normosmics) based on the performance at *SCENTinel 1*.*0*. **B**. Feature importance based on the LDA model across the three smell groups.

## Appendix I

### Eligibility Survey

Thank you for taking the time to determine your eligibility for the validation of our smell test. Smell loss is one of the best predictors of COVID-19 and at present, rapid smell tests that can help monitor the spread of COVID-19 are not available. We have developed our 2-minute smell test for this purpose.

We need your help to make sure that it is accurate and reliable.

We just have a few simple questions to determine your eligibility for this study. Please specify your sex.

□ Male
□ Female
□ Other: _____________
□ Prefer not to answer

Do you currently reside in the United States?

□ Yes
□ No

I am between 18-75 years old.

□ Yes
□ No

Do you currently have a confirmed or diagnosed smell or taste disorder?

□ Yes
□ No

If yes, please specify:

□ Anosmia (complete loss of smell)
□ Hyposmia (partial loss of smell)
□ Parosmia (the quality of some odors has changed)
□ Fluctuations (sometimes I can smell, sometimes I
□ cannot)
□ Other: _______________________

I have access to a smart device (e.g., cell phone, computer, etc.).

□ Yes
□ No

-------------------------------------------------------------------------------------------------------------------------------

You are not eligible to participate in the study.

We immensely appreciate your willingness to help us validate our Rapid Smell Test used for COVID-19 surveillance.

Thank you for your time.

-------------------------------------------------------------------------------------------------------------------------------

Thank you for your time.

You are eligible to participate in the study!

We immensely appreciate your willingness to help us validate our Rapid Smell Test used for COVID-19 surveillance.

For this study, you will receive in the coming day/weeks an envelope with the testing materials.

You will be asked to complete our Rapid Smell Test, and/or another validated smell test, the NIH toolbox, as well as answer some questions about you and your sense of smell online.

Overall your participation will take max ∼10-15 minutes. We ask you to kindly complete the test by September 3rd, 2020.

Do you wish to enroll in the study?

□ Yes
□ No

Please provide your mailing address to receive the smell testing materials (First and Last Name, Street, City, State, ZIP).__________________________________________

I would like for Monell to keep my mailing address so that I receive the Center’s correspondence.

□ Yes
□ No

Are you confident that you would be able to complete the smell test prior to Sept 3rd, 2020? It will only take ∼10-15 minutes maximum.

□ Yes
□ No

The following question is completely optional and will not be used to determine eligibility. Since the intended use of this smell test is to provide a surveillance tool for COVID-19, it is important that we compare it against COVID-19 diagnostic test results (both positive and negative).

(Optional) Would you be willing/have access to receive a COVID-19 nasal-swab test in the next couple of weeks (with or without the presence of symptoms)?

□ Yes
□ No
□ Unsure

Thank you for taking the time to complete our survey.

You will receive in the coming weeks mail from Monell with the testing material.

Stay tuned!

## Notes

### Competing Interest Statement

The authors have declared no competing interest.

### Funding Statement

We acknowledge support from the Monell Institutional Funds. Mackenzie Hannum and Robert Pellegrino are supported by NIH T32 funding (DC000014). This work was also supported by contributions for anosmia research from John Hickey, Philip Johnson, John Labows, Stephen Manheimer, Robert Margolskee, and hundreds of other individual donors. We thank Kerry and the Vernekoff Foundation for their support of this work.

### Author Declarations

The study was approved using a waiver of documentation of consent by the Institutional Review Board of the University of Pennsylvania (#821887).

